# Clinical outcomes and prognostic factors of low-grade serous ovarian cancer: A single-centre observational retrospective study

**DOI:** 10.64898/2026.04.17.26351112

**Authors:** Rewati Prakash, Asifuzzaman Khan, Lawrie Shahbazian, Andrew Arthur, Gabriel Levin, Lucy Gilbert, Carlos M. Telleria

## Abstract

**Objective:** The purpose of the present study is to describe the survival outcomes of patients with low-grade serous ovarian cancer (LGSOC) in the post-operative setting from a tertiary gynecologic oncology referral centre in Quebec, including evaluation of patient characteristics, clinical outcomes and prognostic factors.

**Methods:** The study included 25 patients: 1) with a post-surgical histopathologic diagnosis of a low-grade serous tumour of the ovary, 2) underwent primary cytoreductive surgery prior to adjuvant therapy, and 3) for whom clinical data was available. Clinical and demographic features were characterized by descriptive statistics. Clinical endpoints of progression-free survival (PFS) and overall survival (OS) were assessed, utilizing the Kaplan-Meier method for estimating survival probabilities.

**Results:** The median age of this cohort was 61 years (range, 26-81). Median OS was 140.6 months in patients with no residual disease (R0), 71 months in patients with microscopic residual disease (R1), and 27.7 months in patients with macroscopic residual disease (R2) (p=.001). Residual disease was also found to significantly impact PFS (p=.008). Administration of adjuvant chemotherapy failed to improve survival outcomes altogether (PFS, p = .270; OS, p = .300).

**Conclusions:** This study supports the shifting consensus that optimal cytoreductive surgery, where feasible, is paramount for successful treatment of LGSOC. Furthermore, treatment with adjuvant chemotherapy may lead to worse survival outcomes.

## 1. Introduction

Low-grade serous ovarian cancer (LGSOC) is a rare histopathological subtype of epithelial ovarian cancer, representing 2% of cases all cases in the United States [1]. While traditionally serous ovarian cancer grading was defined on the continuum of tumour cellular atypia, in 2004, LGSOC was reclassified by a two-tier univariate system based on the assessment of cytologic atypia [2]. This system further emphasized that LGSOC is a distinct disease with respect to differing clinical, histological, and molecular characteristics [3].

The origin of LGSOC is still debated, with the majority of research supporting the idea that LGSOC arises in a stepwise fashion, from tubal derived ovarian epithelial inclusions or endosalpingiosis, into serous borderline tumours (SBOTs), to a non-invasive micropapillary SBOT, and then finally to an invasive carcinoma [4–6]. The LGSOC molecular profile is highly distinct from that of high-grade serous ovarian cancer (HGSOC), namely with LGSOC being overwhelmingly TP53 wild type and considered to have comparatively greater genomic stability [7]. Moreover, driver mutations of LGSOC include members of the mitogen activated protein kinase (MAPK) pathway. Multiple studies have demonstrated mutations of KRAS in 16-44% of cases, BRAF in 2-20%, and NRAS in 26% [8, 9]. Interestingly, patients with mutations to these driver genes have greater survival rates [8, 10]. Compared to HGSOC, LGSOC is an indolent malignancy with greater long-term survival and generally diagnosed at a younger age [11–13]. Nevertheless, between 49-80% of patients with LGSOC experience a relapse of disease, usually within 5 years of diagnosis, and at that stage, the disease is incurable [11, 14, 15].

As with other rare malignancies, evaluating the relative efficacy of different treatment strategies is challenging. In the absence of randomized trial data, clinical practice often varies substantially, limiting the development of evidence-based treatment guidelines. An audit of 134 LGSOC patients treated in five different provinces in Canada concluded that there are notable differences in management and outcomes across Canada [16].

In light of the paucity of data and the rarity of the diagnosis, we aim to evaluate the survival outcomes of LGSOC patients in a single tertiary gynecologic oncology referral centre in Quebec.

## 2. Methods

### 2.1 Study Design

A retrospective cohort study at the McGill University and Health Centre (MUHC) Gynecology Oncology Division from January 2000 to March 2025.

### 2.2 Study Population

We included LGSOC who were diagnosed, treated and followed up at the MUHC. Inclusion criteria were 1) age ≥ 18 years; 2) patients having primary surgery within the study period with a surgical pathology diagnosis of LGSOC; 3) patients having surgery and adjuvant treatment and follow-up at MUHC. Exclusion criteria were 1) a non-primary diagnosis of LGSOC, including patients referred to our center after treatment was initiated elsewhere or came for a second option; 2) inoperable tumour; 3) patients returning to referring hospital for treatment and/or follow-up. Details on patient attrition for this study can be found in ***Supplementary Figure 1***.

### 2.3 Data collection

We abstracted data from electronic medical records and scanned documents. Patient demographics collected include age at surgery, parity, body mass index (BMI) as defined by the World Health Organization (WHO) [17], estrogen and progesterone receptor (ER, PR) status, residual disease (RD) status after primary cytoreductive surgery, and the 2014 International Federation of Obstetrics and Gynecology (FIGO) ovarian cancer staging. We defined stages I and II as “early” and stages III and IV were grouped as “late”. Residual disease status was defined according to the following categories: R0 – no visible disease; R1 – less than 1cm; R2 - residual disease of 1cm or greater.

### 2.4 Statistical Analysis

Patient characteristics were presented using absolute and relative frequencies for categorical variables and medians (with minimum and maximum ranges) for continuous variables. The two clinical endpoints measured in this study were progression-free survival (PFS) and overall survival (OS). PFS was defined as the time elapsed from the date of primary cytoreductive surgery to disease progression, recurrence (based on clinical and/or imaging criteria), or death, whichever came first. OS was defined as the time elapsed from the date of a patients’ primary cytoreductive surgery to death from any cause. Patients who did not progress or remained alive were censored at their final tumour evaluation by the MUHC gynecology, radiology, or medical oncology divisions. Survival probabilities were estimated via the Kaplan–Meier method and the Greenwood formula was used to calculate 95% confidence intervals (95% CI). Comparisons of the survival curves were conducted using the Mantel-Cox log-rank test. For all tests, p-values < 0.05 were considered statistically significant. All statistical analysis was conducted with GraphPad Prism 10.5 software (Dotmatics, Boston, MA, USA).

### 2.5 Ethical approval

The study was approved by the institutional ethics review board (MUHC-2022-8522). All patient records were de-identified before inclusion into the study database.

## 3. Results

### 3.1 Patient Characteristics

A total of twenty-five patients were included in the analysis. Detailed patient demographics and clinical characteristics can be found in **Table 1**. The median age at surgery was 61 (range, 26–81) years and the median BMI was 27.3 (range, 16.0–38.9 kg/m^2^). There were ten nulliparous (40%) and fifteen multiparous (60%) women. According to the FIGO staging system, most patients were diagnosed with stage III disease (72%), while 12% and 8% were diagnosed at stages I and II, respectively; 8% had stage IV disease.

**Table 1:** Demographic & Clinical Characteristics of LGSOC Cohort.

| Patient Demographics | All Patients (n=25)* |
| --- | --- |
| Median age at surgery, in years (range) | 61 y (26-81) |
| Median BMI (kg/m<sup>2</sup>) (range) | 27.3 (16.0-38.9) |
| Parity n (%) |  |
| Nulliparous | 10 (40%) |
| Multiparous | 15 (60%) |
| FIGO stage, n (%) |  |
| I | 3 (12%) |
| II | 2 (8%) |
| III | 18 (72%) |
| IV | 2 (8%) |
| Residual Disease after primary surgery, n (%) |  |
| No visible disease; R0 | 11 (44%) |
| Microscopic residual disease; R1<sup>†</sup> | 2 (8%) |
| Macroscopic residual disease; R2<sup>‡</sup> | 7 (28%) |
| Not Reported | 5 (20%) |
| ER Status, n (%) |  |
| Positive | 22 (88%) |
| Negative | 0 |
| Not Tested | 3 (12%) |
| PR Status, n (%) (n=24) |  |
| Positive | 17 (71%) |
| Negative | 3 (13%) |
| Not Tested | 4 (17%) |

BMI – Body Mass Index; FIGO = International Federation of Obstetrics and Gynecology; ER = Estrogen Receptor; PR = Progesterone Receptor
\* – Except where noted otherwise
<sup>†</sup> – Less than 1 cm
<sup>‡</sup> – 1–2 cm: $n = 2$ ; greater than 2 cm: $n = 5$

Following primary cytoreductive surgery, no visible residual disease (R0) was achieved in eleven patients (44%), while two patients (8%) had microscopic residual disease (R1; <1 cm) and seven (28%) had macroscopic residual disease (R2). Among those with R2 disease, 2 patients had residual nodules between 1–2 cm, and 5 patients had residual disease >2 cm.

Hormone receptor testing revealed ER positivity in 22 patients (88%). PR expression was positive in 17 cases (71%).

### 3.2 Systemic Therapy

The majority of the patients (14, 56%) received adjuvant therapy, the details of which are presented in **Table 2**. Eleven patients (44%) received platinum-based chemotherapy (carboplatin and/or paclitaxel); two patients received anti-vascular endothelial growth factor (VEGF) treatment (bevacizumab) in conjunction with chemotherapy, and as maintenance, five patients received anti-hormone therapy as maintenance following chemotherapy. Three patients (12%) received anti-hormone therapy alone as their first-line treatment. Amongst patients who received anti-hormone therapy, the regimens included anastrozole, exemestane, letrozole, and/or tamoxifen. Eleven patients (44%) received no further treatment.

**Table 2:** First-line adjuvant treatments.

| Treatment | Patients (n=25) |
| --- | --- |
| Platinum-based Doublet chemotherapy | 11 (44.0%) |
| Anti-Hormone therapy & Chemotherapy | 5 (45.5%) |
| Anti-VEGF therapy & Chemotherapy | 2 (18.2%) |
| Anti-Hormone therapy | 3 (12.0%) |
| No Treatment | 11 (44.0%) |

VEGF = vascular endothelial growth factor

### 3.3 Progression-Free Survival

The median follow up was 35.6 months (range, 0.3-165.8 months). Sixteen patients (64.0%) experienced a progression of disease, and the median PFS was 34.3 months (95% CI 28.0-99.6) (**Figure 1A**). The median PFS in patients with no residual disease was 78.2 months (95% CI 28.7 – not reached, NR), in R1 it was 67 months (95% CI 28.0 – NR) and in R2 it was 9.6 months (95% CI 0.5 – NR) (p = .008) (**Figure 1B)**. As seen in **Figure 1C**, the median PFS was considerably lower amongst patients diagnosed with late FIGO stage disease (33.8 months, 95% CI 9.6–52.9) as compared to patients diagnosed with early FIGO stage disease (median survival not reached). According to the log rank test, the survival difference was marginally statistically insignificant (p = .063). Nonetheless, this result is indicative of FIGO staging’s high clinical relevance with respect to time-to-progression. Finally, with respect to adjuvant chemotherapy there was no significant difference in median PFS between patients who received no chemotherapy following surgery (52.9 months, 95% CI 5.7 – NR), compared to those who received the chemotherapy (31.8 months, 95% CI 7.1 – 78.2) (p = .270) (**Figure 1D**).

**Figure 1.**
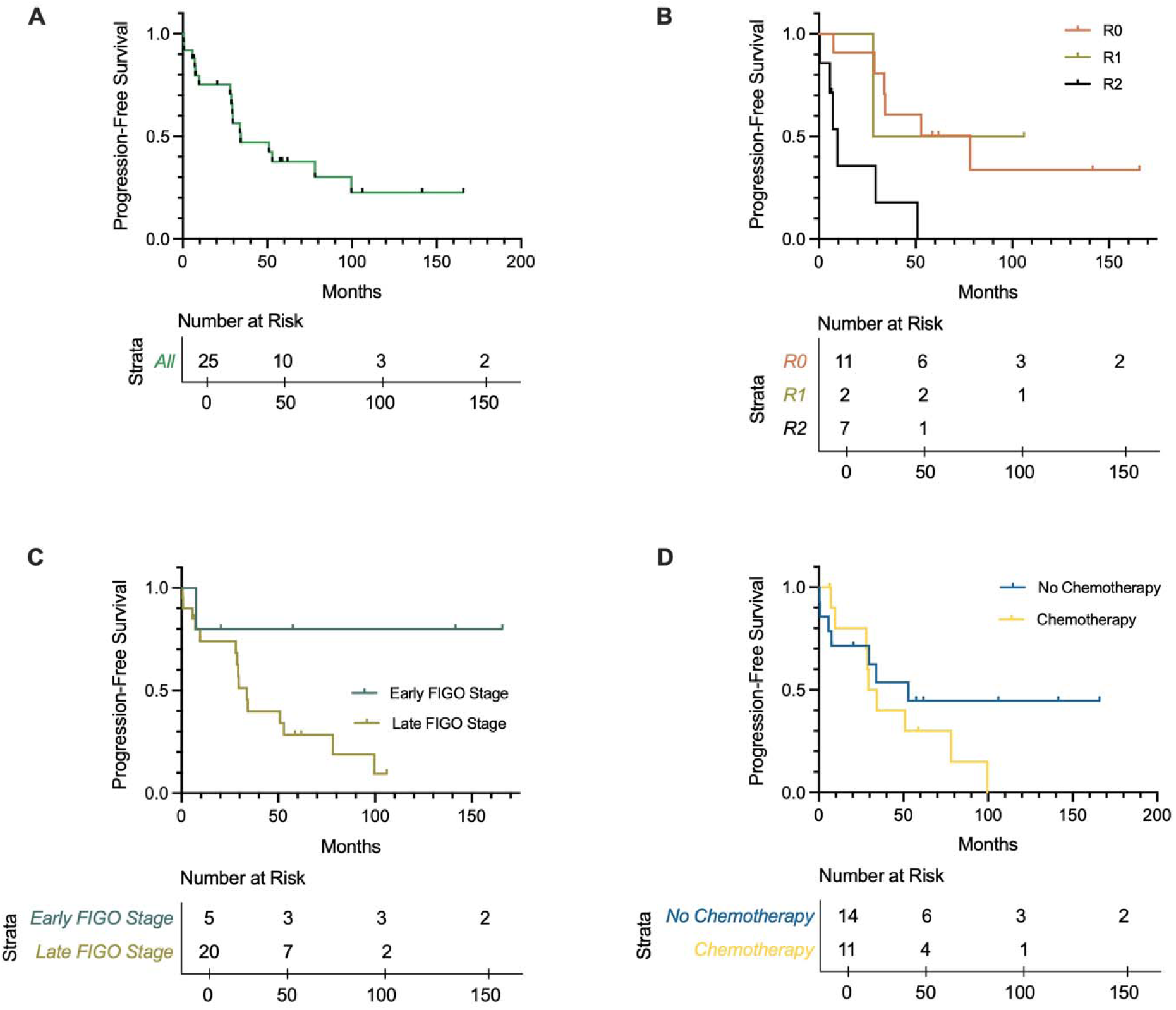
Kaplan-Meier progression-free survival estimates; overall (A), stratified by residual disease after cytoreductive surgery (B), stratified by International Federation of Gynaecology and Obstetrics (FIGO) stages (C), and stratified by first-line treatment with platinum-based chemotherapy (D).

### 3.4 Overall Survival

During follow up, 13 patients died (52.0%). The median OS was 93.0 months (95% CI 29.6–150.3) (**Figure 2A**). The median OS in patients with R0 was 140.6 months (95% CI 31.0 – NE) versus patients with R1 of 71.0 months (95% CI 36 – NE) and patients with R2, 27.7 months (95% CI 4.1 – NE) (p = .001) (**Figure 2B**). Comparing patients based on FIGO staging, the median OS was substantially lower in patients who were diagnosed with late FIGO stage disease (78.2 months, 95% CI 27.7 – 140.6) as opposed to patients diagnosed with early FIGO stage disease (median survival not reached) (**Figure 2C**). This survival difference was again shown to be marginally statistically insignificant (p = .090). Stratifying OS for adjuvant chemotherapy demonstrated a greater median OS in patients who received no chemotherapy (140.6 months, 95% CI 7.4 – NR) than patients who received the adjuvant chemotherapy (78.2 months, 95% CI 27.7 – NR) (p = .30) (**Figure 2D**).

**Figure 2.**
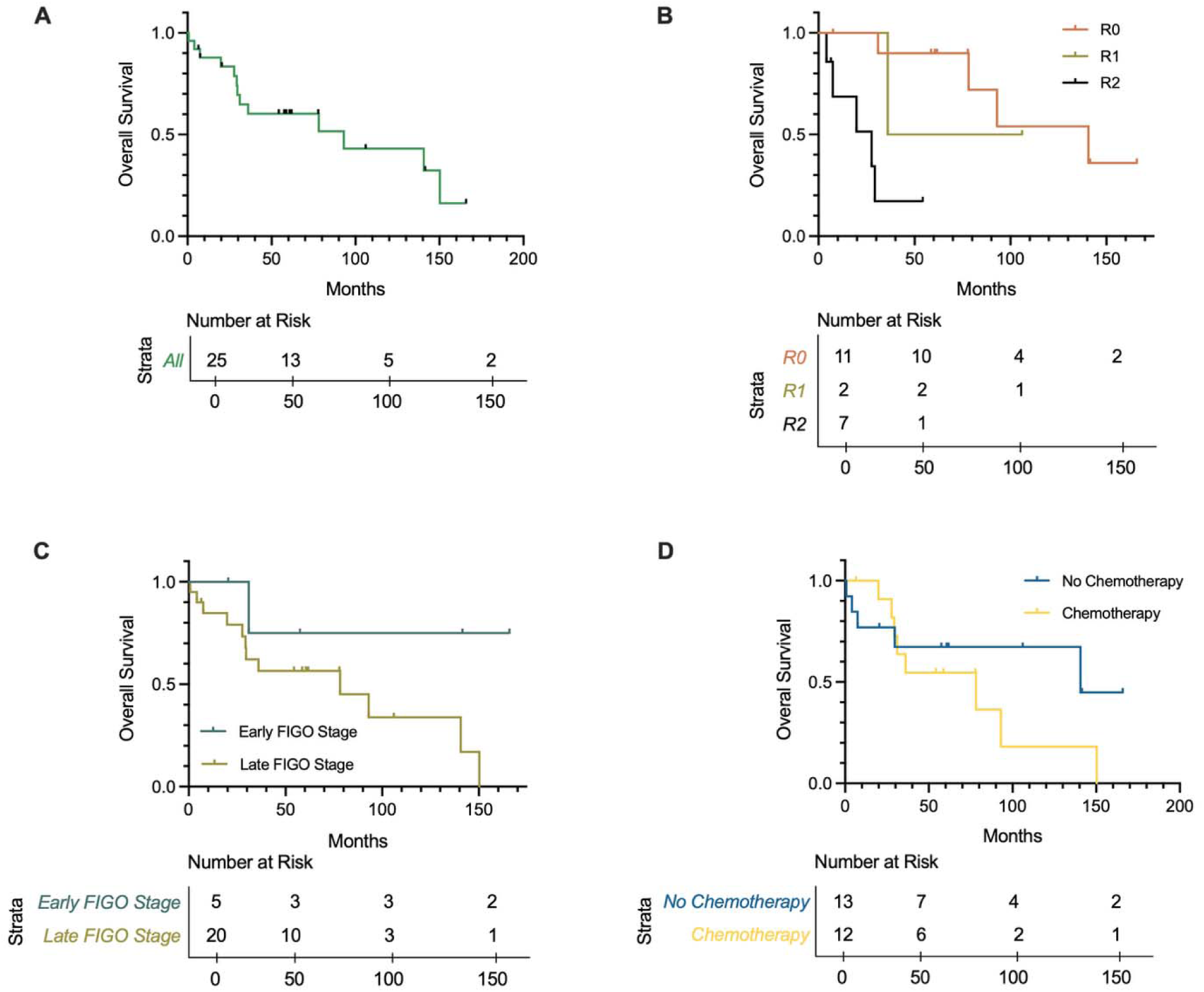
Kaplan-Meier overall survival estimates; overall (A), stratified by residual disease after cytoreductive surgery (B), stratified by International Federation of Gynaecology and Obstetrics (FIGO) stages (C), and stratified by treatment with platinum-based chemotherapy (D).

## 4. Discussion

In this single-centre study of LGSOC patients, the median PFS was 34.3 months, and the median OS was 93.0 months, with residual disease after primary surgery being a strong prognostic factor for both PFS and OS.

LGSOC is generally diagnosed at a median age of 43 years; patients diagnosed under 35 years of age experience the worst outcomes [11, 12]. For decades, the gold standard of treatment for all subtypes of ovarian cancer has included platinum-based chemotherapy. However, many studies have reported significant chemoresistance from LGSOC tumours; as first-line therapy, the objective response rate has been reported as 4-35% and a 2-22% response rate following platinum re-challenge [18–20]. Due to the increased expression of ER and PR in LGSOC, the use of anti-hormone therapy has also become common. Gershenson et al., demonstrated that LGSOC patients following primary cytoreductive surgery who received hormone maintenance therapy alongside chemotherapy were found to have significantly greater survival compared to those who did not [21]. Ongoing research into targeted therapy has focused on the elevated expression of MAPK driver mutations as a therapeutic target for LGSOC tumours. [22, 23].

We found that residual disease status following primary cytoreductive surgery was found to be the most significant predictor of poor prognosis; women with macroscopic residual disease experienced significantly lower survival times compared to women with no residual disease (OS, 27.7 months vs. 140.6 months). These results corroborate multiple prior studies on LGSOC. Di Lorenzo et al., reported that in patients with advanced-stage LGSOC, RD > 10 mm had an OS of 35.2 months, versus 86.4 months in patients with RD 1-10 mm, and 142.3 months in patients with no RD (p = .002) [24]. An analysis of 391 LGSOC cases from the Ovarian Cancer Association Consortium (OCAC) database demonstrated a HR of 2.53 between patients with visible residual disease vs. no visible (p < .001) [25]. However, Gershenson et al., in a 2015 follow up to their 2006 study, reported a lack of significance in median survival when corrected for residual disease; patients with no gross residual disease had a median OS of 98.8 months versus 95.2 months for patients with gross residual disease (p = 0.2) [11]. This lack of association between primary cytoreduction and survival was observed again in a 2024 cohort study that examined the association between residual disease status and survival of less-common epithelial ovarian cancers, which included 1103 cases of LGSOC [26]. Therefore, while the results of our study corroborate the stance of complete surgical resection as a cornerstone for improving survival in LGSOC, other prognostic factors are to be considered.

In the present study, FIGO status at diagnosis and treatment with adjuvant chemotherapy were clinical factors affecting survival in LGSOC. As demonstrated in prior studies, stage at diagnosis was found to have a moderately detrimental effect on prognosis, particularly for PFS, where patients diagnosed at later stages experienced a shorter survival time (33.8 months vs. median survival not reached) [11, 25, 27].

Our data demonstrates that the administration of adjuvant chemotherapy did not significantly improve survival outcomes. The role of systemic chemotherapy treatment for LGSOC is controversial and other studies have likewise indicated a lack of survival benefit with its use. In the aforementioned OCAC database study, the authors did not find any survival benefit from the administration of primary platinum-based chemotherapy (HR 4.59) [25]. Gockley et al., similarly assessed the role of adjuvant chemotherapy treatment in LGSOC patients and found no survival benefit. In a propensity score-matched cohort, the median OS was 95.9 months for patients who did not receive chemotherapy, whereas the median OS was 88.2 months for those who did receive chemotherapy [28]. In a recent prospective study analyzing the effect of adjuvant chemotherapy on LGSOC Johnson et al., found that there was no significant survival difference between patients who were administered adjuvant chemotherapy and those who were not [29]. Although in our study patients only received adjuvant chemotherapy, neoadjuvant chemotherapy (NACT) is often utilized where optimal cytoreduction is not possible. Di Lorenzo et al., found that the response rate to neoadjuvant chemotherapy was 36%, whereas most studies generally report a much lower response to NACT [18, 24]. Nevertheless, patients who received NACT had shorter PFS and OS, on average [24]. Intriguingly, in a 2023 retrospective study assessing responses of LGSOC to neoadjuvant and adjuvant chemotherapy found that 58% of patients following a suboptimal cytoreductive surgery achieved an objective (partial or complete) response to adjuvant treatment while only 1% of patients who received NACT achieved a partial response [18].

Although anti-hormone treatment did not significantly impact survival outcomes in this study, the elevated expression of ER (88%) and PR (71%), suggests a benefit for inclusion of this treatment class. In 2017, women with LGSOC who had previously been treated with adjuvant chemotherapy were additionally treated with hormonal maintenance therapy or continued to be observed. Median PFS was found to be greater in those who had persistent disease and were treated with hormonal maintenance therapy (38.1 months) compared to patients under clinical surveillance alone (15.2 months). The impact of anti-hormonal therapy was even greater when comparing median PFS in patients who were disease free and received the treatment and those who were under clinical surveillance (81.1 vs. 30.0 months) [21]. Furthermore, in the PARAGON study, a phase II trial, found that 61% of patients with ER-positive LGSOC achieved a clinical benefit following treatment with anastrozole for at least 6 months [30].

The main limitations of the present study include the small sample size and the retrospective design. The limited sample size increases the risk of type II errors. Furthermore, the sample size encapsulated insufficient events to prepare an adjusted Cox regression. Our models were thus not able to account for inequivalent distribution of key prognostic factors between each strata of the univariate predictors in each Kaplan-Meier curve. Therefore, the results of the univariate models may be prone to confounder bias. Finally, the inherent flaws of the retrospective design increase the risk of missing or incomplete data, selection bias, and recall bias.

## 5. Conclusion

Overall, while the traditional paradigm for treating ovarian cancer includes the use of systemic chemotherapy, our study supports the idea that treatment of LGSOC with chemotherapy may lead to worse survival outcomes. Our results reinforce the crucial importance of leaving no visible residual disease at primary cytoreductive surgery.

## Data Availability

All data produced in the present study are available upon reasonable request to the authors

## Author Contributions

**Rewati Prakash:** Writing – Original draft preparation, Formal Analysis **Asifuzzaman Khan:** Data Curation, Conceptualization, Writing – Review & Editing **Lawrie Shahbazian:** Conceptualization, Writing – Review & Editing **Andrew Arthur:** Conceptualization, Writing – Review & Editing **Gabriel Levin:** Writing – Review & Editing **Lucy Gilbert:** Conceptualization, Resources, Supervision, Writing – Review & Editing **Carlos M. Telleria:** Supervision, Writing – Review & Editing.

---

**Supplementary Figure 1.**
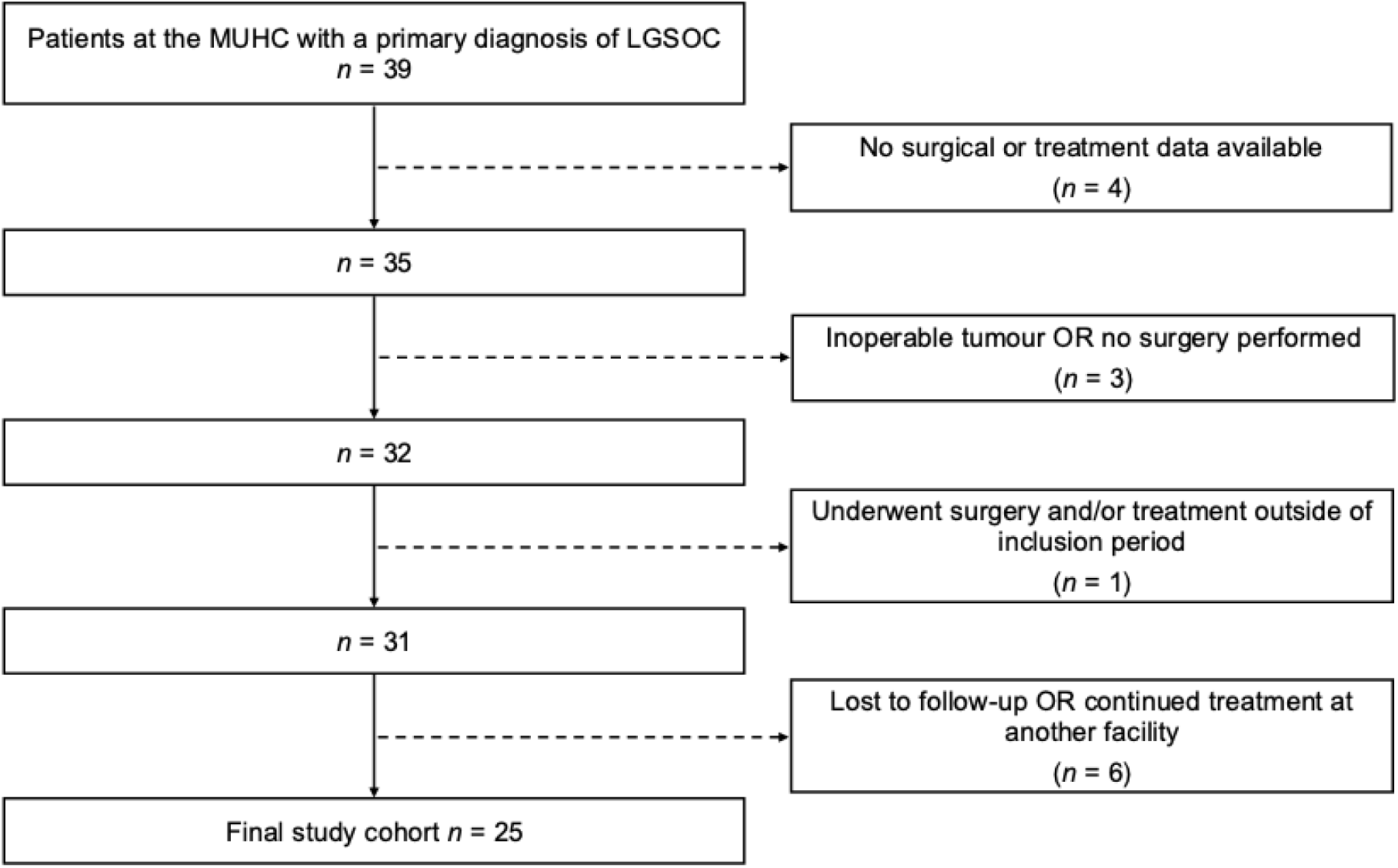
Attrition flow chart identifying patients included in the final study population. LGSOC = low-grade serous ovarian cancer; MUHC = McGill university health centre.

